# Technical Skill Assessment in Minimally Invasive Surgery Using Artificial Intelligence: A Systematic Review

**DOI:** 10.1101/2022.11.08.22282058

**Authors:** Romina Pedrett, Pietro Mascagni, Guido Beldi, Nicolas Padoy, Joël L. Lavanchy

## Abstract

**Objective:** To review artificial intelligence (AI) based applications for the assessment of technical skills in minimally invasive surgery.

**Background:** As technical skill assessment in surgery relies on expert opinion, it is time-consuming, costly, and often lacks objectivity. Analysis of routinely generated data by AI methods has the potential for automatic technical skill assessment in minimally invasive surgery.

**Methods:** A systematic search of Medline, Embase, Web of Science and IEEE Xplore was performed to identify original articles reporting the use of AI in the assessment of technical skill in minimally invasive surgery. Risk of bias (RoB) and quality of included studies were analyzed according to Quality Assessment of Diagnostic Accuracy Studies criteria and the modified Joanna Briggs Institute checklists, respectively. Findings were reported according to the Preferred Reporting Items for Systematic Reviews and Meta-Analyses statement.

**Results:** In total, 1467 articles were identified, 37 articles met eligibility criteria and were analyzed. Motion data extracted from surgical videos (49%) or kinematic data from robotic systems or sensors (46%) were the most frequent input data for AI. Most studies used deep learning (73%) and predicted technical skills using an ordinal assessment scale (73%) with good accuracies in simulated settings. However, all proposed models were in development stage, only 8% were externally validated and 16% showed a low RoB.

**Conclusion:** AI is promising to automate technical skill assessment in minimally invasive surgery. However, models should be benchmarked on representative datasets using predefined performance metrics and tested in clinical implementation studies.

**Mini Abstract:** Technical skill assessment in minimally invasive surgery is time consuming and costly. Artificial intelligence is a promising technology to facilitate and automate technical skill assessment. Therefore, this article systematically reviews artificial intelligence applications for the assessment of technical skills in minimally invasive surgery.

## INTRODUCTION

The assessment of technical skill is of major importance in surgical education and quality improvement programs given the association of technical skills with clinical outcomes^1–4^. This correlation has been demonstrated amongst others in bariactric^1^, upper gastrointestinal^2^ and colorectal surgery^3,4^. In addition, data from the American Colleges of Surgeons National Surgical Quality Improvement Program revealed that surgeon’s technical skills as assessed by peers during right hemicolectomy are correlated with outcomes in colorectal as well as in non-colorectal surgeries performed by the same surgeon^3^, showing the overarching impact of technical skills on surgical outcomes.

To date, technical skills are assessed through direct observations of surgeons’ performance or retrospectively by reviewing surgical video recordings. Generally, this process involves either classifying skill levels in ordinal scales (e.g., novice, intermediate and expert) through unstructured observations or assessing performance intervals through the use of structured, validated checklists (e.g., Objective Structured Assessment of Technical Skills (OSATS)^5^, Global Evaluative Assessment of Robotic Skills (GEARS)^6^ (Figure 1). Therefore, technical skill assessment is complex and time consuming, hence costly. Moreover, technical skill assessment is limited by inter-observer variability and reviewer bias^7^.

**Figure 1:**
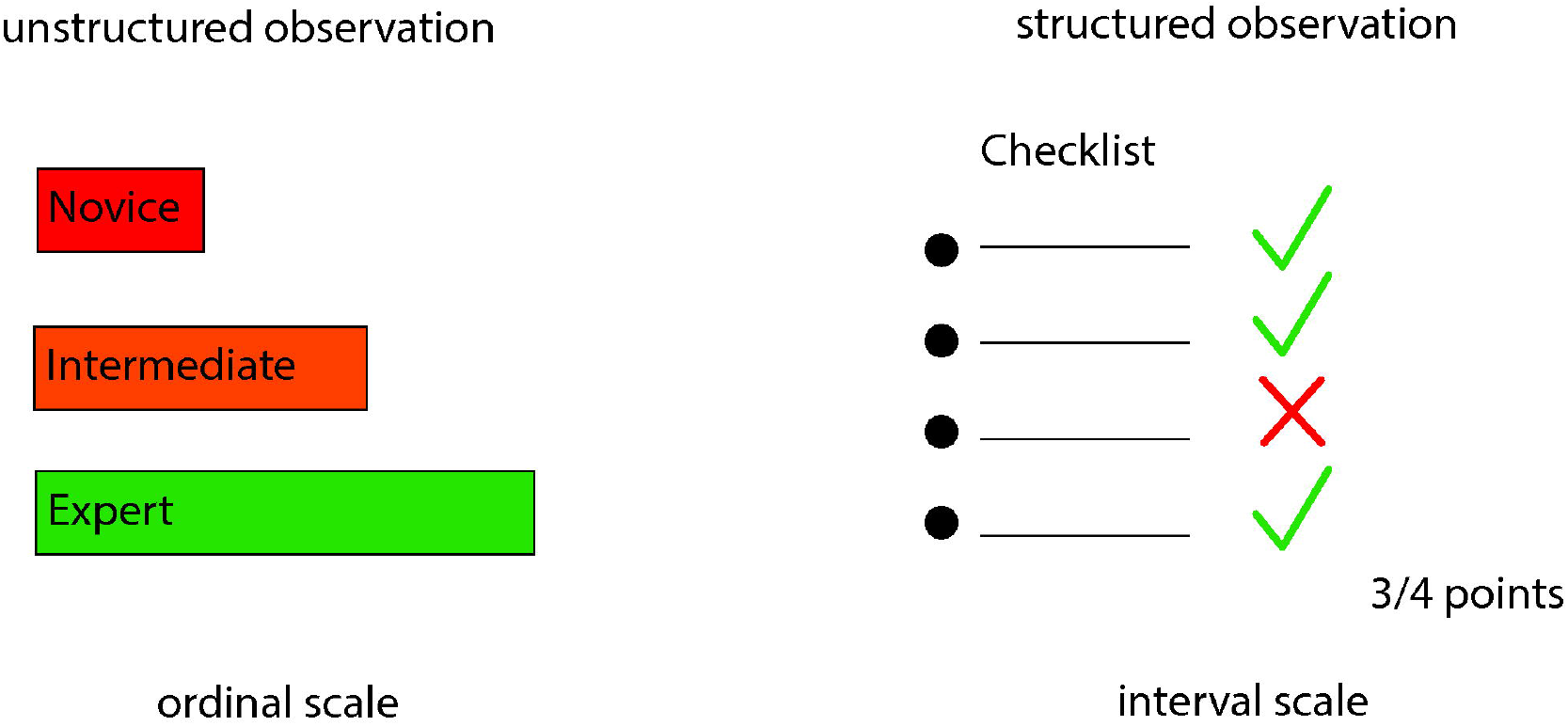
Human technical skill assessment in minimally invasive surgery.

The growing adoption of minimally invasive surgery and recent developments in artificial intelligence (AI) could lead to automatic, objective, and consistent technical skill assessment in surgery.

As in minimally invasive surgery the surgical field is visualized by cameras, surgical videos are easily recorded and readily available at a large scale. Surgical videos can be used to extract information about technical skills. In robotic surgery the movements of the surgeon are translated to robotic arms holding the endoscope and the instruments. This allows the extraction of kinematic data such as moving trajectories directly form the robotic system. Based on kinematic data performance metrics of technical skills were developed^8^.

AI is a very promising technology that is widely adopted in medicine^9,10^. For example, AI is able to detect diabetic retinopathy^11,12^ and to screen for lung cancer^13^ and malignant skin cancer^14^ with an accuracy comparable to expert clinician screening.

Two subfields of AI are particularly used to extract and analyze motion data from surgical videos or robotic systems: machine learning (ML) and deep learning (DL). ML can be defined as computer algorithms that learn distinct features iterating over data without explicit programming. DL designates computer algorithms that analyze unstructured data using neural networks (NN). NN are computer algorithms designed in analogy to the synaptic network of the human brain. The input data is processed through multiple interconnected layers of artificial neurons, each performing mathematical operations on the input data to predict an output. The predicted output is compared to the human labeled output to optimize the operations of the NN, which makes it a self-learning system. From an AI perspective technical skill assessment is a classification (prediction of expert levels) or a regression task (prediction of a score). Figure 2 illustrates how different input data types are processed by AI models to predict technical skills.

**Figure 2:**
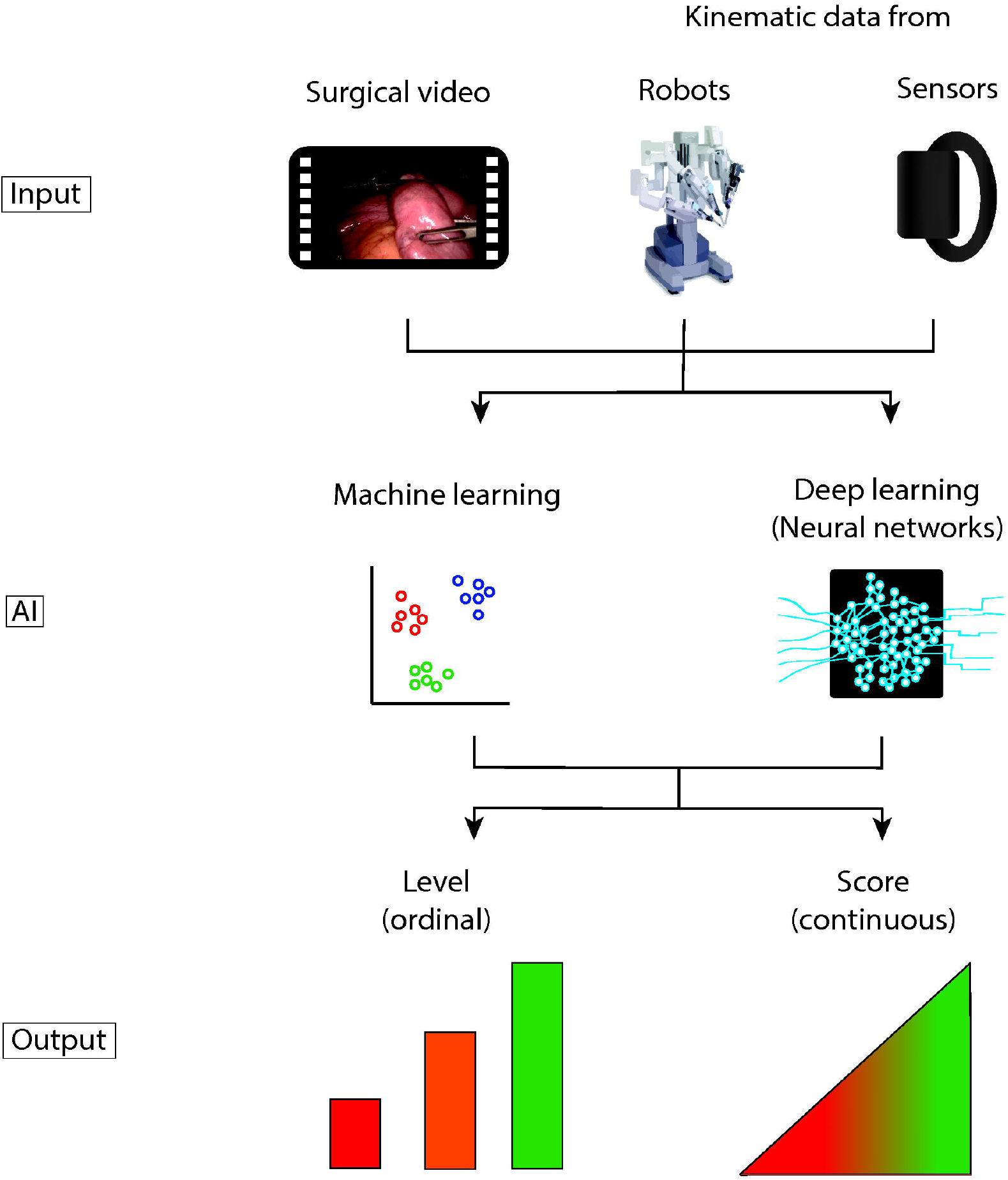
Automated technical skill assessment in minimally invasive surgery by artificial intelligence.

The aim of this systematic review was to analyze studies using AI for technical skill assessment in minimally invasive surgery.

## METHODS

This systematic review is reported in accordance with the PRISMA (Preferred Reporting Items for Systematic Reviews and Meta-Analyses)^15^ guidelines and was prospectively registered at PROSPERO (2021 CRD42021267714).

### Literature search

A systematic literature search of the databases Medline/Ovid, Embase/Ovid, Web of Science and IEEE Explore was conducted on August 25^th^, 2021. The first three databases account for biomedical literature and IEEE Explore for technical literature. A librarian of the University Library, University of Bern performed the literature search combining the following terms using Boolean operators: 1) Minimally invasive surgery including endoscopic, laparoscopic, or robotic surgery, and box model trainer. 2) AI including machine learning, supervised learning, unsupervised learning, computer vision and convolutional neural networks. 3) Technical skill assessment including surgical skill assessment, surgical performance assessment, and task performance analysis. The full-text search terms can be found in the Supplementary. The literature search was re-run prior to final analysis on February 25^th^, 2022.

### Eligibility criteria

Studies presenting original research on AI applications for technical skills assessment in minimally invasive surgery including box model trainers published within the last 5 years (08/2016-02/2022) in English language were included. Review articles, conference abstracts, comments, and letters to the editor were excluded.

### Study selection

Before screening, the identified records were automatically deduplicated using the reference manager program Endnote™ (Clarivate Analytics). After removal of the duplicates, two authors (R.P. & J.L.L.) independently screened the titles and abstracts of the identified records for inclusion using the web-tool Rayyan (https://www.rayyan.ai)^16^. Disagreement of the two authors regarding study selection was settled in joint discussion. Of all included records the full-text articles were acquired. Articles not fulfilling the inclusion criteria after full-text screening were excluded.

### Data extraction

Besides bibliographic data (title, author, publication year, journal name), the study population, the setting (laparoscopic/robotic simulation or surgery), the task assessed (e.g., peg transfer, cutting, knot-tying), the data input (motion data from video recordings, kinematic data from robotic systems or sensors), the dataset used, the assessment scale (ordinal scale vs. interval scale), the AI models used (ML or DL), the performance and the maturity level (development, validation, implementation) of AI models were extracted from the included studies.

### Performance metrics

Performance metrics included accuracy, precision, recall, F1-score, and Area Under the Curve of Receiver Operator Characteristic (AUC-ROC). Accuracy is the proportion of correct predictions among the total number of observations. Precision is the proportion of true positive predictions among all (true and false) positive predictions and referred to as the positive predictive value. Recall is the proportion of true positive predictions among all relevant observations (true positives and false negatives) and referred to as sensitivity. F1-score is the harmonic mean of precision and recall and is a measure of model performance. A ROC curve plots the true positive against the false positive predictions at various thresholds and the AUC describes performance of the model to distinguish true positive from false positive predictions.

### Risk of bias and quality assessment

The risk of bias of the included studies was assessed using the modified version of Quality Assessment of Diagnostic Accuracy Studies (QUADAS-2) criteria^17^. The quality of studies was evaluated using the modified Joanna Briggs Institute critical appraisal checklist for cross-sectional research in ML as used in^18,19^.

## RESULTS

The literature search retrieved a total of 1467 studies. After removing all duplicates, the remaining 1236 studies were screened by title and abstract. Thereafter, 88 studies remained, of which 51 were excluded after full-text screening. In summary, 37 studies^20–56^ met eligibility criteria and thus were included into this systematic review (Figure 3). Out of these 37 studies, six (16%)^27,35,41,47,52,54^ were obtained during the re-run prior to final analysis six months after the initial literature search was conducted. Table 1 gives an overview on the 37 studies included in this systematic review (for full information extracted see Supplementary Table S1).

**Table 1:** Information summary of all studies included in this review

| Author | Year | Population | Setting | Tasks | Input Data | Dataset | Assessment | AI model | Accuracy | Maturity level |
| --- | --- | --- | --- | --- | --- | --- | --- | --- | --- | --- |
| Alonso-Silverio et al.<sup>20</sup> | 2018 | 20 | LS | PC | VR | private | binary (experienced, non-experienced) | DL | 0.94 | dev |
| Anh et al.<sup>21</sup> | 2020 | 8 | RS | SU, NP, KT | KD (dV) | JIGSAWS | N, I, E | DL | 0.97 | dev |
| Baghdadi et al.<sup>22</sup> | 2018 | na | Lap | Pelvic lymph node dissection | VR | private | PLACE score | DL | 0.83 | dev |
| Benmansour et al.<sup>23</sup> | 2018 | 6 | RS | SU, NP, KT | KD (dV) | JIGSAWS | Custom score | DL | na | dev |
| Brown et al.<sup>24</sup> | 2017 | 38 | RS | PT | KD (dV) | private | GEARS score (1-5: exact rating) | ML | 0.75 | dev |
| Castro et al.<sup>25</sup> | 2019 | 8 | RS | SU, NP, KT | KD (dV) | JIGSAWS | N, I, E | DL | 0.98 | dev |
| Fard et al.<sup>26</sup> | 2017 | 8 | RS | SU, KT | KD (dV) | JIGSAWS | binary (N, E) | ML | 0.9 | dev |
| Fathabadi et al.<sup>27</sup> | 2021 | na | LS | PC | VR | private | Level A (excellent) - E (very bad) | DL | na | dev |
| Fawaz et al.<sup>28</sup> | 2019 | 8 | RS | SU, NP, KT | KD (dV) | JIGSAWS | N, I, E | DL | 1 | dev |
| Forestier et al.<sup>29</sup> | 2018 | 8 | RS | SU, NP, KT | KD (dV) | JIGSAWS | N, I, E | ML | 0.96 | dev |
| French et al.<sup>30</sup> | 2017 | 98 | LS | PT, SU, PC | VR | private | binary (N, E) | ML | 0.9 | dev |
| Funke et al.<sup>31</sup> | 2019 | 8 | RS | SU | VR | JIGSAWS | N, I, E | DL | 1 | dev |
| Gao et al.<sup>32</sup> | 2020 | 13 | LS | PC | fNIRS data | private | FLS score: pass/fail | DL | 0.91 | dev |
| Islam et al.<sup>33</sup> | 2016 | 52 | LS | PT, SU, PC | VR | private | Custom score | DL | na | dev |
| Jin et al.<sup>34</sup> | 2018 | na | Lap | Lap cholecystectomy | VR | m2cai16-tools-location | unknown | DL | na | dev |
| Juarez-Villalobos et al.<sup>35</sup> | 2021 | 8 | RS | SU, NP, KT | KD (dV) | JIGSAWS | binary (N, E) | ML | 1 | dev |
| Keles et al.<sup>36</sup> | 2021 | 33 | LS | PT, threading | fNIRS data | private | binary (student vs. attending) | ML | ~ 0.9 | dev |
| Kelly et al.<sup>37</sup> | 2020 | na | LS | PT, SU, PC, Clipping | VR | private | binary (N, E) | DL | 0.97 | dev |
| Khalid et al.<sup>38</sup> | 2020 | 8 | RS | SU, NP, KT | VR | JIGSAWS | N, I, E | DL | 0.77 | dev |
| Kitaguchi et al.<sup>39</sup> | 2021 | na | Lap | Lap colorectal surgery | VR | private | ESSQS score | DL | 0.75 | dev |
| Kowalewski et al.<sup>40</sup> | 2019 | 28 | LS | SU, KT | KD (s) | private | N, I, E | DL | 0.7 | dev |
| Lajkó et al.<sup>41</sup> | 2021 | 8 | RS | SU, NP, KT | KD (dV) | JIGSAWS | binary (N, E) | DL | 0.84 | dev |
| Lavanchy et al.<sup>42</sup> | 2021 | 40 | Lap | Lap cholecystectomy | VR | private | binary (good vs. poor)<br>5-point Likert scale (+/- 1 point) | ML, DL | 0.87<br>0.7 | dev |
| Laverde et al.<sup>43</sup> | 2018 | 7 | LS | PT | KD (s) | private | N, I, E | DL | na | dev |
| Law et al.<sup>44</sup> | 2017 | 12 | Rob | Robotic prostatectomy | VR | private | binary (good vs. poor) | ML, DL | 0.92 | dev |
| Lee et al.<sup>45</sup> | 2020 | 1/ na | RS, Rob | Robotic thyroid surgery/simulation | VR | private | N, I, E | DL | 0.83 | dev |
| Liu et al.<sup>46</sup> | 2020 | na | Lap | Lap gastrectomy | VR | private | modified OSATS score | DL | na | dev |
| Liu et al.<sup>47</sup> | 2021 | 8 / na | RS, Lap | SU, NP, KT / Lap surgery gastric cancer | VR | JIGSAWS | modified OSATS score | ML | na | dev |
| Lyman et al.<sup>48</sup> | 2021 | 2 | RS | SU of hepaticojejunostomy | KD (dV) | private | binary (N, I) | ML | 0.89 | dev |
| Nguyen et al.<sup>49</sup> | 2019 | 8 | RS | SU, NP, KT | KD (dV) | JIGSAWS | N, I, E | DL | 0.98 | dev |
| Oquendo et al.<sup>50</sup> | 2018 | 32 | LS | SU | KD (s) | private | OSATS scores (+/- 4 points) | ML | 0.89 | dev |
| Pérez-Escamirosa et al.<sup>51</sup> | 2019 | 43 | LS | PT, SU, PC | VR | private | binary (experienced vs. non-experienced) | ML | 0.98 | dev |
| Soleymani et al.<sup>52</sup> | 2021 | 8 | RS | SU, NP, KT | VR | JIGSAWS | N, I, E | DL | 0.97 | dev |
| Uemura et al.<sup>53</sup> | 2018 | 67 | LS | SU | KD (s) | private | binary (N, E) | DL | 0.79 | dev |
| Wang Y. et al.<sup>54</sup> | 2021 | 18 | RS | SU | VR | private | N, I, E<br>GEARS score (+/- 1 point) | DL | 0.83<br>0.86 | dev |
| Wang Z. et al.<sup>55</sup> | 2018 | 8 | RS | SU, NP, KT | KD (dV) | JIGSAWS | N, I, E | DL | 0.95 | dev |
| Wang Z. et al.<sup>56</sup> | 2018 | 8 | RS | SU, NP, KT | KD (dV) | JIGSAWS | N, I, E | DL | 0.96 | dev |

**Figure 3:**
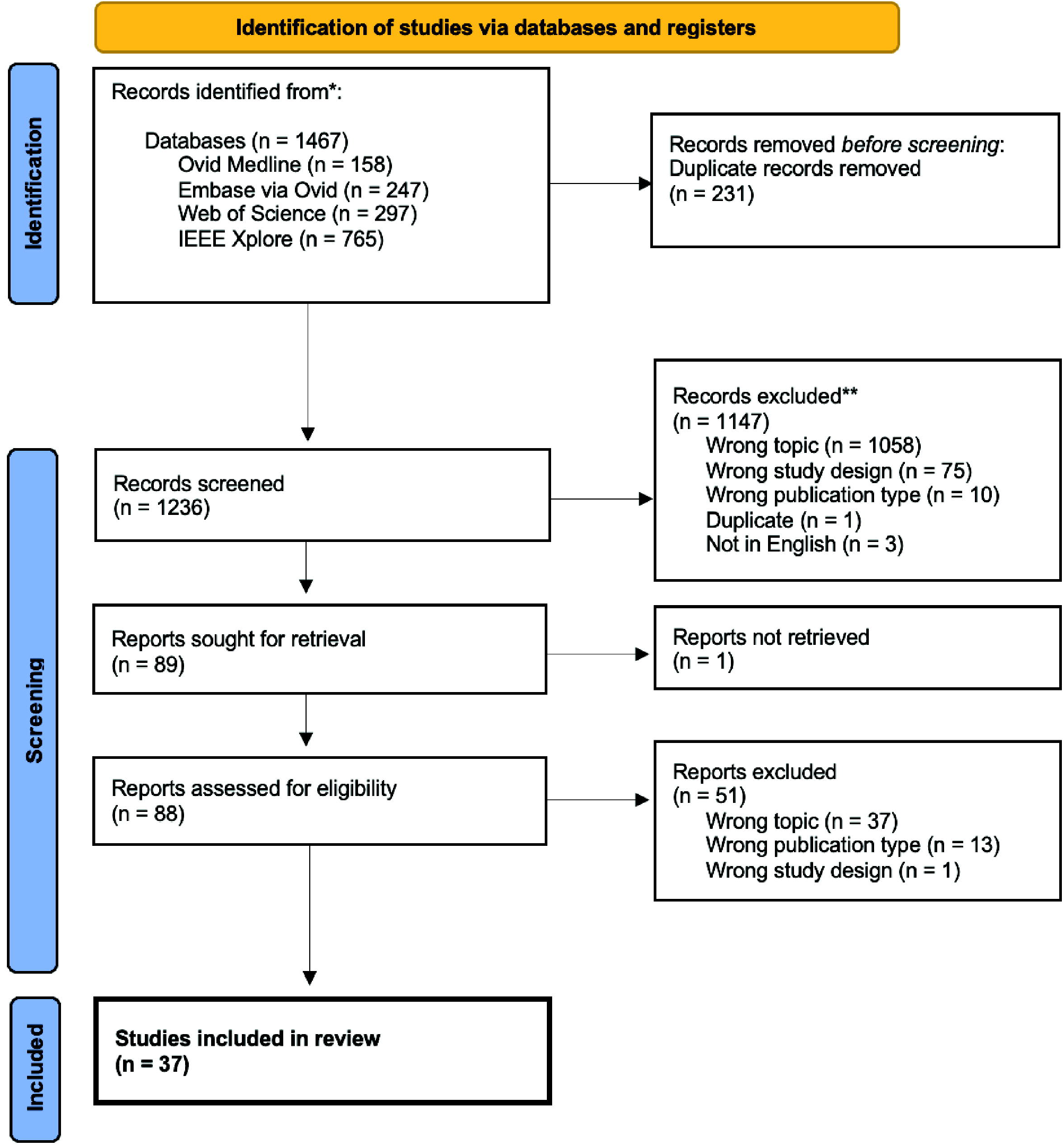
PRISMA flow diagram of the study selection process (from PRISMA Statement 2020^15^).

### Settings and tasks

Most often, motion data from surgical videos or kinematic data from robotic systems or sensors were collected from simulators rather than during actual surgical procedures. The most common simulators used were robotic box models (51%, n=19) ^21,23–26,28,29,31,35,38,41,45,47–49,52,54–56^. Laparoscopic simulators were the second most common setting for data collection (32%, n=12)^20,27,30,32,33,36,37,40,43,50,51,53^.

The most common tasks assessed were suturing (62%, n=23)^21,23,25,26,28–31,33,35,37,38,40,41,47,49–56^, knot-tying (41%, n=15) ^21,23,25,26,28,29,35,38,40,41,47,49,52,55,56^ and needle passing (35%, n=13) ^21,23,25,28,29,35,38,41,47,49,52,55,56^. Other tasks assessed were peg transfers (19%, n=7)^24,30,33,36,37,43,51^ and pattern cutting (19%, n=7)^20,27,30,32,33,37,51^. All of these tasks are part of the Fundamentals of Laparoscopic Surgery program, a well-established and validated training curriculum for laparoscopic surgery^57,58^.

Eight studies (22%)^22,34,39,42,44–47^ used data of real surgical procedures. Six (16%)^22,34,39,42,46,47^ of them using videos of laparoscopic surgeries as for example laparoscopic cholecystectomies^34,42^ or laparoscopic pelvic lymph node dissections^22^. Two studies (5%)^44,45^ used video data obtained from robotic surgeries such as robotic prostatectomy^44^ or robotic thyroid surgery^45^. The tasks assessed in surgical procedures ranged from entire interventions to specific steps (e.g., lymph node dissection^22^, clip application^42^).

### Input data and datasets

Three different types of input data were used throughout the 37 studies: video data (49%, n=18)^20,22,27,30,31,33,34,37–39,42,44–47,51,52,54^, kinematic data (46%, n=17)^21,23–26,28,29,35,40,41,43,48–50,53,55,56^ and functional near-infrared spectroscopy (fNIRS) data (5%, n=2)^32,36^. Video recordings either from endoscopic laparoscopic/robotic camera or external cameras are used in 18 studies (49%). Kinematic data was obtained from DaVinci robotic systems (Intuitive Surgical Inc., CA, USA) in 13 studies (35%)^21,23–26,28,29,35,41,48,49,55,56^ and from external sensors in four studies (11%)^40,43,50,53^. For example, electromyography sensors (Myo armband, Thalmic Labs, Ontario, CA)^40^, optical sensors (Apple Watch, Apple, CA, USA)^43^ or magnetic sensors attached to the instruments^50,53^ were used as external sensors to collect kinematic data. Two studies^32,36^ recorded fNIRS data from participants while they performed laparoscopic tasks. For example, Keles et al.^36^ collected fNIRS data using a wireless, high density NIRS device, measuring functional brain activation of the prefrontal cortex. The NIRS device was adjacent to the surgeons’ foreheads while they performed different laparoscopic tasks.

Publicly available datasets were used in 16 studies (43%)^21,23,25,26,28,29,31,34,35,38,41,47,49,52,55,56^. Of those, the JIGSAWS (Johns Hopkins University and Intuitive Surgical, Inc. Gesture and Skill Assessment Working Set)^59^ dataset was most frequently used (n=15, 41%)^21,23,25,26,28,29,31,35,38,41,47,49,52,55,56^. It contains video and kinematic data together with human annotated skill ratings of eight surgeons performing three surgical tasks in five-fold repetition in a robotic box model trainer. One study^34^ extended the publicly available m2cai16-tool dataset^60^ with locations of surgical tools and published it as m2cai16-tools-localisation dataset. Though, most studies (n=21, 57%) created private datasets, that were not publicly released. Most datasets (n=34, 92%) were monocentric. However, three studies (8%) used a multicentric dataset: French, et al.^30^ used a multi-institutional dataset from three centers, Kitagutchi, et al.^39^ draw a sample form a national Japan Society of Endoscopic Surgeons database, and Law, et al.^44^ used a part of a statewide national quality improvement database collected by the Michigan Urological Surgical Improvement Collaborative. Three of the 37 studies included (8%)^29,47,49^, reported external validation on a second independent dataset.

### Assessment

Technical surgical skills can be assessed using expert levels (ordinal scale) or proficiency scores (interval scale) (Figure 1). In 27 of the studies (73%) an ordinal scale was applied^20,21,25–31,35–38,40–45,48,49,51–56^. In twelve studies (32%) participants were categorized in two different skill levels^20,26,30,35–37,41,42,44,48,51,53^ and in 14 studies (38%) into three different expert levels (novice, intermediate, expert)^21,25,28,29,31,38,40,43,45,49,52,54–56^. Ten studies (27%) applied different proficiency scores: Pelvic Lymphadenectomy Assessment and Completion Evaluation (PLACE^61^), Fundamentals of Laparoscopic Surgery (FLS^62^), Endoscopic Surgical Skill Qualification System (ESSQS^63^), Objective Structured Assessment of Technical Skills (OSATS^64^), and Global Evaluative Assessment of Robotic Skills (GEARS^6^)^22–24,32,33,39,46,47,50,54^.

### AI models

All AI models in this review are either ML- or DL-based. ML was applied in 12 studies (32%)^24,26,29,30,35,36,40,42,47,48,50,51^ and DL in 27 studies (73%)^20–23,25,27,28,31–34,37–46,49,52–56^. Two studies (5%) used a combination of ML and DL models^42,44^.

### Performance

The most common performance metrics reported in the studies included in this systematic review is accuracy (n=30, 81%)^20–22,24–26,28–32,35–42,44,45,48–56^. Accuracies of the best performing models range between 0.7 – 1. Other performance metrics reported include F1-score (n=6, 16%)^31,35,38,43,50,51^, recall (n=4, 11%)^24,31,38,40^, sensitivity (n=4, 11%)^20,32,50,51^, specificity (n=4, 11%)^20,32,50,51^, and AUC-ROC (n=4, 11%)^20,35,50,51^. Four studies (11%)^23,27,33,34^ did not report a performance metric at all.

### Risk of bias and quality assessment

Six of the included studies (16%)^24,37,39,40,42,50^ had an overall low probability of bias in the risk of bias assessment. The other studies had one (n=10, 27%), two (n=9, 24%), three (n=8, 22%), four (n=3, 8%) or five criteria (n=1, 3%) at risk of bias. The full risk of bias assessment table is presented in the Supplementary (Table S2). The quality assessment of the included studies is displayed in Figure 4. All proposed AI models were in a developmental preclinical stage of maturity, none was implemented in routine clinical use.

**Figure 4:**
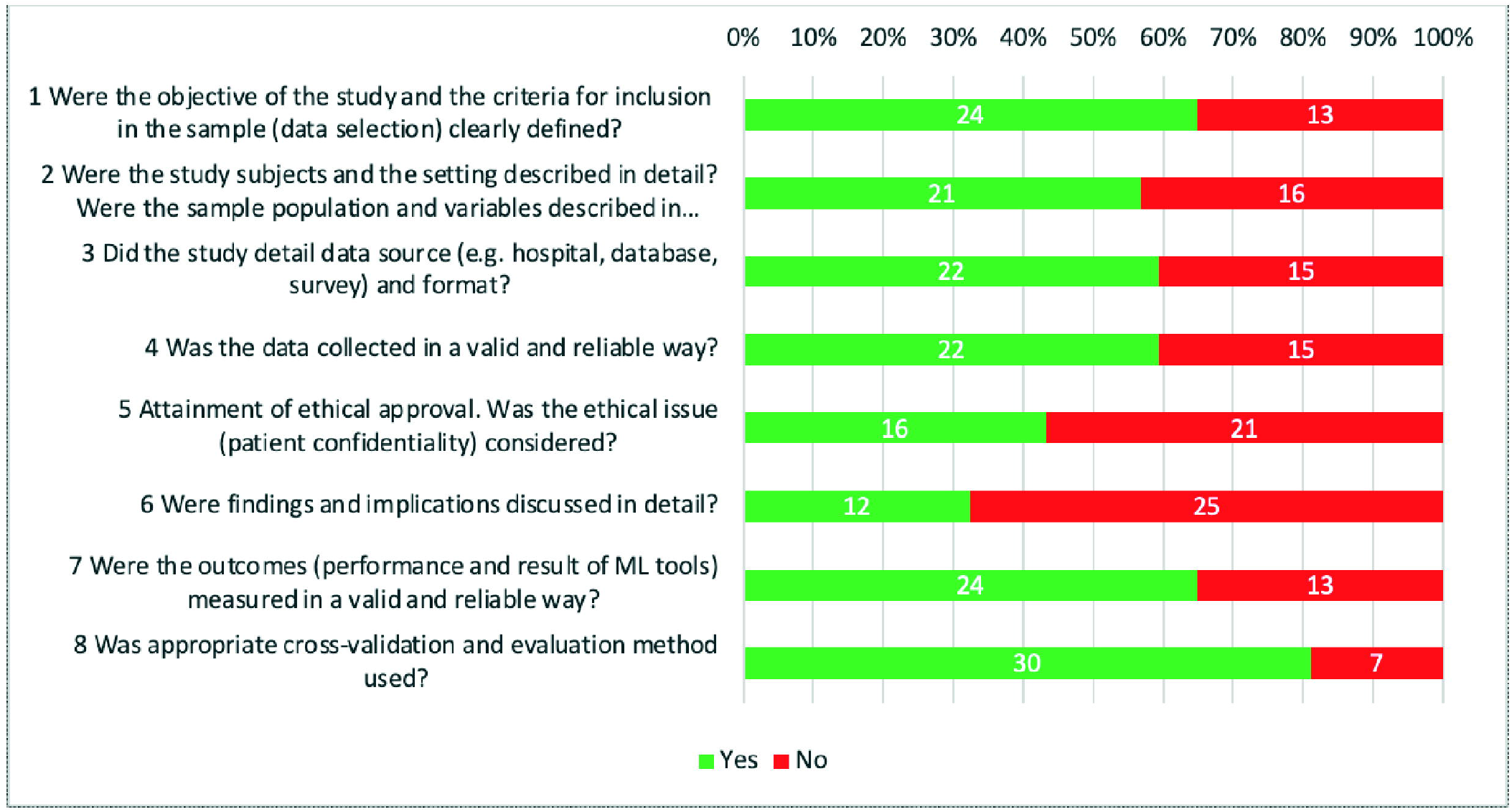
Quality assessment of the included studies. The numbers within the bars represent the respective number of studies.

## DISCUSSION

This systematic review of AI applications for technical skill assessment in minimally invasive surgery included 37 studies. Technical surgical skills were assessed in 51% of studies in robotic simulators, in 32% of studies in laparoscopic simulators, and in 22% of studies in actual surgical procedures. The input data to AI models were video data (49%), kinematic data from robotic systems or sensors (46%), and fNIRS data (2%). Technical skills were classified in 73% of studies using skill levels and in 27% of studies using proficiency scores. In total, 32% of AI models were ML-based and 73% of AI models were DL-based. Most studies (81%) reported accuracy as performance metric. Overall, 84% of studies were at risk of bias and only 3 studies tested their AI model for external validity using a second independent dataset. None of the proposed models was implemented in routine clinical use.

The comparability of studies included in this systematic review is limited due to several fundamental differences between them. Most studies (57%) use private datasets of different settings, tasks, and sizes. However, 15 studies included in this systematic review used JIGSAWS, a robotic simulator dataset and the most frequently used dataset in technical skill assessment. The use of simulators for technical skill assessment has advantages and disadvantages. On the one hand, simulators allow to control the experimental setting and enable reproducibility of studies. On the other hand, box model trainers simulate surgical tasks and have only a restricted degree of realism. In addition, simulators are well established in surgical training but have limited significance in the assessment of fully trained surgeons. The use of video recordings and motion data of actual surgeries as input data improves the validity of technical skill assessment models. However, in actual surgeries the experimental setting cannot be standardized and therefore, lacks reproducibility.

Moreover, the comparison of studies is impaired by the different scales and scores used to measure technical skill. Some studies use ordinal scales with different numbers of skill levels (good vs. bad^20,26,35– 37,41,42,44,48,51,53^, novice vs. intermediate vs. expert^21,25,28–31,38,40,43,45,49,52,54–56^) others use different interval scales (OSATS scores^28,40,50^, GEARS scores^24,54^, or Likert scales^42^). This finding represents the general difficulty to define and measure technical surgical skills.

Most of the studies included in this systematic review have methodologic limitations. Overall, 84% of studies included in this review are at risk of bias. The quality assessment of the included studies revealed that only 32% of studies discussed the findings and implications in detail. Furthermore, only three studies included in this review have a multicentric dataset. Only three of the AI models studied are validated on an independent external dataset. Therefore, it is questionable whether the AI models included in this review would generalize to other settings, tasks, and institutions. Out of 37 included studies, 30 report on accuracy. However, there is a large variation of reported performance metrics among studies included in this systematic review. Due to the novelty of AI application in the healthcare domain and in surgery in particular, the literature lacks standards in the evaluation of AI methods and their performance. There is an urgent need for the application of guidelines to assess AI models and for studies comparing them head-to-head. Guidelines for early stage clinical evaluation of AI^65^ and clinical trials involving AI^66^ have been published recently. However, the studies included in this review are all at a preclinical stage where these guidelines do not apply. A multi-stakeholder initiative recently introduced guidelines and flowcharts on the choice of AI evaluation metrics in the medical image domain^67^. For surgical video analysis this effort still needs to be taken^68^. To overcome the limitations of the proposed AI models for technical skill assessment, valid and representative datasets using predefined performance metrics, and external validation in clinical implementation studies will be essential.

Looking at the educational benefits of AI algorithms, the current models allow an estimation of individual skill levels in comparison with the population the algorithm was trained on. However, no direct or concrete feedback on how to improve technical skills is provided. Potentially training AI models on assessment scores divided in different domains of technical skills (e.g. bimanual dexterity, tissue handling,) could help to give automated but actionable feedback.

In conclusion, AI has great potential to automate technical skill assessment in minimally invasive surgery. Various AI models, that analyze surgical video or movement data from simulators or actual surgical procedures and correlate them with technical surgical skills, have been studied. However, the studies included in this review lack standardization of datasets, performance metrics and external validation. Therefore, we advocate for benchmarking of AI models on valid and representative datasets using predefined performance metrics and testing in clinical implementation studies.

## Data Availability

All data produced in the present work are contained in the manuscript.

## ACKNOLEDGEMENTS

We would like to acknowledge the help of Tanya Karrer, Information Specialist Medicine, University Library, University of Bern with the literature search.

