## Supplementary for "Technical Skill Assessment in Minimally Invasive Surgery Using Artificial Intelligence: A Systematic Review"

### Full-text search terms of the search strategy

Definitive Literature Search Strategy conducted by Tanya Karrer, Information Specialist Medicine, Research Support Services, University Library, University of Bern, on August 25th 2021 and re-run on February 25th 2022.

**OVID MEDLINE**(R) and Epub Ahead of Print, In-Process, In-Data-Review & Other Non-Indexed Citations, Daily and Versions(R) < 1946 to August 24, 2021 > . 25.8.2021

1 exp Minimally Invasive Surgical Procedures/ 551322

2 Robotic Surgical Procedures/ 10673

3 Surgery, Computer-Assisted/ 18834

4 Specialties, Surgical/ 4298

5 Surgeons/ed [Education] 1043

6 (Laparoscop* or minimal-invasive surg* or minimally invasive surg* or celioscop* or peritoneoscop* or box-model* or box model* or remote operation* or telerobotic* or tele-robotic* or tele robotic* or robot* surg* or robot-assisted surg* or robot assisted surg* or robot-enhanced surg* or robot enhanced surg* or surgical robot* or surgery robot* or computer-assisted surg* or computer assisted surg* or surg* education).tw. 155871

7 or/1-6 627683

8 artificial intelligence/ or machine learning/ or deep learning/ or supervised machine learning/ or unsupervised machine learning/ or neural networks, computer/ or Pattern Recognition, Automated/ 89828

9 (artificial intelligence or artificial surg* or neural network* or comput* network model* or machine learning or machine-learning or deep learning* or deep-learning or natural language process* or motion tracking* or automat* pattern recognition* or comput* intelligence or machine intelligence or computer reasoning or computational reasoning or computer vision* or computational vision* or machine sensing).tw. 121663

10 8 or 9 162542

11 ((surgical or surgeon* or laparoscopic or clinical) adj6 (skill* or performance or technique* or quality or process* or model* or level*) adj6 (assess* or evaluat* or classification or rating or scor* or accuracy)).tw. 41471

12 Clinical competence/ 98831

13 Professionalism/ed [Education] 163

14 Education, Medical, Continuing/ 25232

15 Laparoscopy/ed [Education] 1794

16 Surgeons/ed [Education] 1043

17 "Task Performance and Analysis"/ 32098

18 11 or 12 or 13 or 14 or 15 or 16 or 17 188918

19 7 and 10 and 18 212

20 limit 19 to last 5 years

**Results: 142**

https://ovidsp.ovid.com/ovidweb.cgi?T=JS&NEWS=N&PAGE=main&SHAREDSEARCHID=6mwmKdYyxRIM7S9MsMe1vGNE323wr8aTObZxUuKBd8V9KCAB4xRFCZZhYcQXls1eE

**EMBASE VIA OVID** <1974 to 2021 August 24 >, 25.8.2021

1 exp minimally invasive surgery/ 46081
2 exp computer assisted surgery/ 24547
3 exp laparoscopic surgery/ 89074
4 exp laparoscopy/ 172635
5 surgical technology/ 2054
6 (Laparoscop* or minimal-invasive surg* or minimally invasive surg* or celioscop* or peritoneoscop* or box-model* or box model* or remote operation* or telerobotic* or tele-robotic* or tele robotic* or robot* surg* or robot-assisted surg* or robot assisted surg* or robot-enhanced surg* or robot enhanced surg* or surgical robot* or surgery robot* or computer-assisted surg* or computer assisted surg* or surg* education).ab,kw,ti. 255614
7 1 or 2 or 3 or 4 or 5 or 6 323807
8 artificial intelligence/ 33832
9 deep learning/ 17462
10 supervised machine learning/ 2224
11 unsupervised machine learning/ 1173
12 machine learning/ or exp artificial neural network/ or automated pattern recognition/ or network learning/ 112821
13 (artificial intelligence or artificial surg* or neural network* or comput* network model* or machine learning or machine-learning or deep learning* or deep-learning or natural language process* or motion tracking* or automat* pattern recognition* or comput* intelligence or machine intelligence or computer reasoning or computational reasoning or computer vision* or computational vision* or machine sensing).ti,ab,kw. 156778
14 8 or 9 or 10 or 11 or 12 or 13 199420
15 clinical competence/ 63852
16 professionalism/ 9841
17 continuing education/ 31781
18 exp surgical training/ 22826
19 task performance/ 147487
20 ((surgical or surgeon* or laparoscopic or clinical) adj6 (skill* or performance or technique* or quality or process* or model* or level*) adj6 (assess* or evaluat* or classification or rating or scor* or accuracy)).ti,ab,kw. 60521
21 15 or 16 or 17 or 18 or 19 or 20 326609
22 7 and 14 and 21 262
23 limit 22 to last 5 years
**Results: 189**

https://ovidsp.ovid.com/ovidweb.cgi?T=JS&NEWS=N&PAGE=main&SHAREDSEARCHID=2va9FFfK1QL 2YjNsjNzGNRax2EXAD4RKfK1aLDyojM2ST9e8mUAeL3Xvnz7xD4tOh

**WEB OF SCIENCE**, 25.8.2021
((TS=(Laparoscop* or minimal-invasive surg* or minimally invasive surg* or celioscop* or peritoneoscop* or box-model* or box model* or remote operation* or telerobotic* or telerobotic* or tele robotic* or robot* surg* or robot-assisted surg* or robot assisted surg* or robotenhanced surg* or robot enhanced surg* or surgical robot* or surgery robot* or computerassisted surg* or computer assisted surg* or surg* education or surg* training or surg* technolog*)) AND TS=(artificial intelligence or artificial surg* or neural network* or comput* network model* or machine learning or machine-learning or deep learning* or deep-learning* or natural language process* or motion tracking* or automat* pattern recognition* or comput* intelligence or machine intelligence or computer reasoning or computational reasoning or computer vision* or computational vision* or machine sensing)) AND TS=((surgical OR surgeon* OR laparoscopic OR clinical) NEAR/6 (skill* OR performance OR technique* OR quality OR process* OR model* OR level*) NEAR/6 (assess* OR evaluat* OR classification OR rating* OR scoring* OR accuracy)) AND DOP=(2016-01-01/2021-12-31)
**Results: 265**

https://www.webofscience.com/wos/woscc/summary/97217178-1812-43fe-a337-6ffa3e14978e-057db63a/relevance/1

**IEEE XPLORE**, 25.8.2021
In all Metadata (Includes the abstract, index terms, and bibliographic citation data (such as document title, publication title, author, etc.) ((((("All Metadata":Laparoscop* OR "All Metadata":"minimally invasive surgery" OR "All Metadata":"minimal-invasive surgery" OR "All Metadata":robot* surg* OR "All Metadata":"remote operation*" OR "All Metadata":"computer assisted surgery" OR "All Metadata":"box-model")))) AND ((("All Metadata":skill* OR "All Metadata":performance OR "All Metadata":technique* OR "All Metadata":quality OR "All Metadata":level OR "All Metadata":assessment OR "All Metadata":evaluation OR "All Metadata":classification OR "All Metadata":rating OR "All Metadata":scoring OR "All Metadata":accuracy)))) AND ((("All Metadata":"artificial intelligence" OR "All Metadata":machine learning OR "All Metadata":"deep learning" OR "All Metadata":"deeplearning" OR "All Metadata":"natural language processing" OR "All Metadata":"computer vision" OR "All Metadata":"machine sensing" OR "All Metadata":"neural network" OR "All Metadata":"neural networks")))
Filter: Conferences, Journals, Early Access Articles, Standards, Years 2016-2021
**Results: 583**

https://ieeexplore.ieee.org/search/searchresult.jsp?contentType=all&refinements=ContentType%3AConferences&refinements=ContentType%3AJournals&refinements=ContentType%3AEarly+Access+Articles&refinements=ContentType%3AStandards&sortType=&ranges=2016_2021_Publication Year&matchBoolean=true&searchField=Search_All&queryText=((((((Search_All:Laparoscop*+OR+Search_All:%22minimally+invasive+surgery%22+OR+Search_All:%22minimal-invasive+surgery%22+OR+Search_All:robot*+surg*+OR+Search_All:%22remote+operation*%22+OR+Search_All:%22computer+assisted+surgery%22+OR+Search_All:%22box-model%22))))+AND+(((Search_All:skill*+OR+Search_All:performance+OR+Search_All:technique*+OR+Search_All:quality+OR+Search_All:level+OR+Search_All:assessment+OR+Search_All:evaluation+OR+Search_All:classification+OR+Search_All:rating+OR+Search_All:scoring+OR+Search_All:accuracy))))+AND+(((Search_All:%22artificial+intelligence%22+OR+Search_All:machine+learning+OR+Search_All:%22deep+learning%22+OR+Search_All:%22deep-learning%22+OR+Search_All:%22natural+language+processing%22+OR+Search_All:%22computer+vision%22+OR+Search_All:%22machine+sensing%22+OR+Search_All:%22neural+network%22+OR+Search_All:%22neural+networks%22))))&history=no

Table S2: Data extracted from the 37 studies included in this systematic review

| Title | Authors | Year | Country | Journal | Population | Setting | Tasks | Input Data | AI model | Assessment | Results | | | | | | | | |
|---|---|---|---|---|---|---|---|---|---|---|---|---|---|---|---|---|---|---|---|
| **Development of a Laparoscopic Box Trainer Based on Open Source Hardware and Artificial Intelligence for Objective Assessment of Surgical Psychomotor Skills[1]** | Gustavo A. Alonso-Silverio, Fernando Pérez-Escamirosa, Raúl Bruno-Sanchez, José L. Ortiz-Simon, Roberto Muñoz-Guerrero, Arturo Minor-Martinez and Antonio Alarcón-Paredes | 2018 | Mexico | Surgical Innovation | 20 volunteers | LS | PC | VR | ANN | binary (experienced, non-experienced) | Validation scheme | Accuracy | Sensitivity | Specificity | AUC | | | | |
| | | | | | | | | | | | Holdout (67/33) | 93.94% | 0.78 | 1 | 0.97 | | | | |
| | | | | | | | | | | | 10-fold cross-validation | 91% | 0.78 | 0.96 | 0.93 | | | | |
| | | | | | | | | | | | Leave one out | 88% | 0.74 | 0.93 | 0.91 | | | | |
| **Towards near real-time assessment of surgical skills: A comparison of feature extraction techniques[2]** | Nguyen Xuan Anh, Ramesh Mark Nataraja, Sunita Chauhan | 2020 | Australia | Computer Methods and Programs in Biomedicine | 8 participants | RS | SU, NP, KT | KD (dV) | DL, CNN | N, I, E | Method | 3s | | 4s | | 5s | | 6s | |
| | | | | | | | | | | | | 1s | 2s | 1s | 2s | 1s | 2s | 1s | 2s |
| | | | | | | | | | | | **Suturing Task: Accuracy (effect of window size)** | | | | | | | | |
| | | | | | | | | | | | CNN | 96.84 | 96.84 | 96.9 | 96.87 | 96.92 | 96.82 | 97.04 | 96.92 |
| | | | | | | | | | | | LSTM | 95.09 | 94.57 | 96.39 | 95.67 | 96.45 | 95.8 | 96.48 | 96.45 |
| | | | | | | | | | | | CNN-LSTM | 96.39 | 96.3 | 96.44 | 96.19 | 96.64 | 96.05 | 96.69 | 96.16 |
| | | | | | | | | | | | Auto-encoder | 83.46 | 79.81 | 80.7 | 79.94 | 81 | 80.69 | 81.04 | 80.96 |
| | | | | | | | | | | | **Needle passing** | | | | | | | | |
| | | | | | | | | | | | CNN | 95.36 | 94.96 | 95.65 | 95.12 | 95.83 | 95.46 | 96.44 | 95.88 |
| | | | | | | | | | | | LSTM | 91.52 | 90.67 | 93.63 | 92.19 | 93.83 | 92.94 | 94.06 | 93.05 |
| | | | | | | | | | | | CNN-LSTM | 93.45 | 92.26 | 93.05 | 92.73 | 93.44 | 93.16 | 94.2 | 94.11 |
| | | | | | | | | | | | Auto-encoder | 82.25 | 80.33 | 80.36 | 80.18 | 80.13 | 79.98 | 81.5 | 80.25 |
| | | | | | | | | | | | **Knot tying** | | | | | | | | |
| | | | | | | | | | | | CNN | 92.75 | 92.16 | 93.02 | 92.94 | 93.13 | 93.51 | 93.56 | 93.49 |
| | | | | | | | | | | | LSTM | 89.57 | 89.38 | 91.31 | 89.88 | 91.41 | 89.94 | 91.56 | 90.36 |
| | | | | | | | | | | | CNN-LSTM | 90.98 | 90.24 | 91.74 | 90.25 | 91.95 | 90.53 | 93.01 | 91.1 |
| | | | | | | | | | | | Auto-encoder | 80.63 | 76.02 | 77.75 | 75.95 | 77.6 | 75.15 | 77.69 | 75.54 |
| **A computer vision technique for automated assessment of surgical performance using surgeons' console-feed videos[3]** | Amir Baghdadi, Ahmed A. Hussein, Youssef Ahmed, Lora A. Cavuoto, Khurshid A. Guru | 2018 | USA | International Journal of Computer Assisted Radiology and Surgery | unknown | Lap | Pelvic lymph node dissection | CR | CV | PLACE (pelvic lymphadenectomy appropriateness and completion evaluation) | Accuracy | 0.833 | | | | | | | |

| | | | | | | | | | | | | | | |
|---|---|---|---|---|---|---|---|---|---|---|---|---|---|---|
| **A neural network architecture for automatic and objective surgical skill assessment[4]** | Malik Benmansour, Wahida Handouzi, Abed Malti | 2018 | Algeria | PROCEEDINGS 2018 3RD INTERNATIONAL CONFERENCE ON ELECTRICAL SCIENCES AND TECHNOLOGIES IN MAGHREB (CISTEM) | 6 participants (2 intermediates, 4 novices) | LS | SU, NP, KT | KD (dV) | DNN | Custom score | not available | | | |
| **Using Contact Forces and Robot Arm Accelerations to Automatically Rate Surgeon Skill at Peg Transfer[5]** | Jeremy D. Brown, Conor E. O'Brien, Sarah C. Leung, Kristoffel R. Dumon, David I. Lee, and Katherine J. Kuchenbecker | 2017 | USA | IEEE transactions on bio-medical engineering | 38 participants (3 trials each) | RS | PT | KD (dV) | ML | GEARS scores (1-5: exact rating) | **Accuracy** | GEARS Domain | Regression Learner | Classification Learner |
| | | | | | | | | | | | | Depth Perception | 63.3 +/- 9.5% | 71.7 +/- 9.5% |
| | | | | | | | | | | | | Bimanual Dexterity | 66.7 +/- 11.8% | 53.3 +/- 16.2% |
| | | | | | | | | | | | | Efficiency | 73.3 +/- 16.0% | 58.3 +/- 8.3% |
| | | | | | | | | | | | | Force Sensitivity | 63.3 +/- 9.5% | 51.7 +/- 10.9% |
| | | | | | | | | | | | | Robotic Control | 71.7 +/- 12.6% | 75.0 +/- 15.6% |
| | | | | | | | | | | | **Precision** | GEARS Domain | | |
| | | | | | | | | | | | Regression | Depth Perception | 0.47 +/- 0.37 | |
| | | | | | | | | | | | | Bimanual Dexterity | 0.53 +/- 0.38 | |
| | | | | | | | | | | | | Efficiency | 0.64 +/- 0.36 | |
| | | | | | | | | | | | | Force Sensitivity | 0.43 +/- 0.39 | |
| | | | | | | | | | | | | Robotic Control | 0.47 +/- 0.40 | |
| | | | | | | | | | | | Classification | Depth Perception | 0.40 +/- 0.40 | |
| | | | | | | | | | | | | Bimanual Dexterity | 0.28 +/- 0.30 | |
| | | | | | | | | | | | | Efficiency | 0.50 +/- 0.38 | |
| | | | | | | | | | | | | Force Sensitivity | 0.26 +/- 0.28 | |
| | | | | | | | | | | | | Robotic Control | 0.44 +/- 0.43 | |
| | | | | | | | | | | | **Recall** | | | |
| | | | | | | | | | | | Regression | Depth Perception | 0.56 +/- 0.38 | |
| | | | | | | | | | | | | Bimanual Dexterity | 0.55 +/- 0.38 | |
| | | | | | | | | | | | | Efficiency | 0.65 +/- 0.36 | |

| | | | | | | | | | | | | | | |
|---|---|---|---|---|---|---|---|---|---|---|---|---|---|---|
| | | | | | | | | | | | | Force Sensitivity | 0.45 +/- 0.41 | |
| | | | | | | | | | | | | Robotic Control | 0.55 +/- 0.44 | |
| | | | | | | | | | | | Classification | Depth Perception | 0.48 +/- 0.47 | |
| | | | | | | | | | | | | Bimanual Dexterity | 0.38 +/- 0.41 | |
| | | | | | | | | | | | | Efficiency | 0.54 +/- 0.33 | |
| | | | | | | | | | | | | Force Sensitivity | 0.37 +/- 0.40 | |
| | | | | | | | | | | | | Robotic Control | 0.51 +/- 0.47 | |
| **Towards Optimizing Convolutional Neural Networks for Robotic Surgery Skill Evaluation**[6] | Dayvid Castro, Danilo Pereira, Cleber Zanchettin, David Macêdo, Byron L. D. Bezerra | 2019 | Brazil | International Joint Conference on Neural Networks | 8 participants | RS | SU, NP, KT | KD (dV) | CNN | N, I, E | | Micro-Accuracy | Makro-Accuracy | |
| | | | | | | | | | | | Suturing Task | 98.37 | 98.65 | |
| | | | | | | | | | | | Needle-passing | 98.89 | 98.98 | |
| | | | | | | | | | | | Knot-tying | 98.9 | 98.68 | |
| **Automated robot-assisted surgical skill evaluation: Predictive analytics approach**[7] | Mahtab J. Fard, Sattar Ameri, R. Darin Ellis, Ratna B. Chinnam, Abhilash K. Pandya, Michael D. Klein | 2017 | USA | The International Journal of Medical Robotics and Computer Assisted Surgery | 8 participants | RS | SU, KT | KD (dV) | ML | binary (N, E) | Accuracy | LOSO (LR best) | LOUO (SVM best) | |
| | | | | | | | | | | | Suturing Task | 89.9 | 79.8 | |
| | | | | | | | | | | | Knot-tying | 82.3 | 77.9 | |
| **Surgical Skill Assessment System Using Fuzzy Logic in a Multi-Class Detection of Laparoscopic Box-Trainer Instruments**[8] | Fatemeh Rashidi Fathabadi, Janos L. Grantner, Saad A Shebrain, Ikhlas Abdel-Qader | 2021 | USA | IEEE International Conference on Systems, Man, and Cybernetics (SMC) | unknown | LS | PC | VR | DL | Levels A (excellent)-E (very bad) | not available | | | |
| **Accurate and interpretable evaluation of surgical skills from kinematic data using fully convolutional neural networks**[9] | Hassan Ismail Fawaz, Germain Forestier, Jonathan Weber, Lhassane Idoumghar, Pierre-Alain Muller | 2019 | France | International Journal of Computer Assisted Radiology and Surgery | 8 participants | RS | SU, NP, KT | KD (dV) | CNN | N, I, E | | Micro-Accuracy | Makro-Accuracy | Spearman's Coefficient |
| | | | | | | | | | | | Suturing Task | 100 | 100 | 0.6 |
| | | | | | | | | | | | Needle-passing | 100 | 100 | 0.57 |
| | | | | | | | | | | | Knot-tying | 92.1 | 93.2 | 0.65 |
| **Surgical motion analysis using discriminative interpretable patterns**[10] | Germain Forestier, François Petitjean, Pavel Senin, Fabien Despinoy, Arnaud Huaulmé, Hassan Ismail Fawaz, Jonathan Weber, Lhassane Idoumghar, Pierre-Alain Muller, Pierre Jannin | 2018 | France | Artificial Intelligence In Medicine | 8 participants | RS | SU, NP, KT | KD (dV) | ML | N, I, E | | Micro-Accuracy/ Precision (LOSO) | Makro-Accuracy / Precision (LOSO) | |
| | | | | | | | | | | | Suturing Task | 89.74 | 86.67 | |
| | | | | | | | | | | | Needle-passing | 96.3 | 95.83 | |
| | | | | | | | | | | | Knot-tying | 61.11 | 53.33 | |

| | | | | | | | | | | | | | | | | |
|---|---|---|---|---|---|---|---|---|---|---|---|---|---|---|---|---|
| **Predicting surgical skill from the first N seconds of a task: value over task time using the isogony principle[11]** | Anna French, Thomas S. Lendvay, Robert M. Sweet, Timothy M. Kowalewski | 2017 | USA | International Journal of Computer Assisted Radiology and Surgery | 98 surgeons | LS | PT, SU, PC | VR | ML | N, E / N, I, E | **Accuracy (2-class classification)** | Peg transfer | Cutting | Suturing | Best % accuracy mean/median | Min time to 90% of best accuracy |
| | | | | | | | | | | | DA | 80.1 (0.13) | 86.1 (0.11) | 83.0 (0.2) | 83.7/85.7 | 2 s |
| | | | | | | | | | | | QDA | 87.6 (0.12) | 86.9 (0.11) | 66.0 (0.24) | 82.5/82.6 | 4 s |
| | | | | | | | | | | | SVM | 85.2 (0.12) | 90.2 (0.10) | 82.6 (0.20) | 87.2/88.7 | 8 s |
| | | | | | | | | | | | LR | 84.6 (0.12) | 90.2 (0.10) | 81.5 (0.21) | 86.6/86.4 | 7 s |
| | | | | | | | | | | | **Accuracy (3-class classification)** | Peg transfer | Cutting | Suturing | Best % accuracy mean/median | Min time to 90% of best accuracy |
| | | | | | | | | | | | LDA = linear discriminant analysis | 57.3 (0.13) | 63.0 (0.14) | 66.0 (0.16) | 61.6/61.7 | 2 s |
| | | | | | | | | | | | QDA = quadratic discriminant analysis | 66.6 (0.12) | 59.2 (0.14) | 44.3 (0.16) | 58.9/58.6 | 4 s |
| | | | | | | | | | | | SVM | 62.2 (0.13) | 67.0 (0.13) | 65.5 (0.14) | 65.1/65.5 | 3 s |
| | | | | | | | | | | | LR = logistic regression | 60.8 (0.12) | 68.3 (0.13) | 65.7 (0.15) | 65.1/64.3 | 3 s |
| **Video-based surgical skill assessment using 3D convolutional neural networks[12]** | Isabel Funke, Sören Torge Mees, Jürgen Weitz, Sefanie Speidel | 2019 | Germany | International Journal of Computer Assisted Radiology and Surgery | 8 participants | RS | SU, NP, KT | VR | DL, CNN | N, I, E | **Knot tying** | Accuracy | Avg. Recall | Avg. F1 | | |
| | | | | | | | | | | | 3D ConvNet (RGB) | 95.8 +/- 1.6 | 95.6 +/- 1.2 | 95.9 +/- 1.5 | | |
| | | | | | | | | | | | 3D ConvNet (OF) | 95.1 +/- 2.7 | 94.2 +/- 3.2 | 95.0 +/- 2.9 | | |
| | | | | | | | | | | | **Suturing** | Accuracy | Avg. Recall | Avg. F1 | | |
| | | | | | | | | | | | 3D ConvNet (RGB) | 100 +/- 0 | 100 +/- 0 | 100 +/- 0 | | |
| | | | | | | | | | | | 3D ConvNet (OF) | 100 +/- 0 | 100 +/- 0 | 100 +/- 0 | | |
| | | | | | | | | | | | **Needle passing** | Accuracy | Avg. Recall | Avg. F1 | | |
| | | | | | | | | | | | 3D ConvNet (RGB) | 96.4 +/- 0 | 96.3 +/- 0 | 96.6 +/- 0 | | |
| | | | | | | | | | | | 3D ConvNet (OF) | 100 +/- 0 | 100 +/- 0 | 100 +/- 0 | | |
| **Functional Brain Imaging Reliably Predicts Bimanual Motor Skill Performance in a Standardized Surgical Task[13]** | Yuanyuan Gao, Pingkun Yan, Uwe Kruger, Lora Cavuoto, Steven Schwaitzberg, Suvranu De, Xavier Intes | 2020 | USA | IEEE transactions on bio-medical engineering | 13 medical students | LS | PC | fNIRS data (functional near-infrared spectroscopy) | DL | FLS (Fundamentals of Laparoscopic Surgery) Score: Pass/fail | | Accuracy | Sensitivity | Specificity | Computation time (ms) | |
| | | | | | | | | | | | Brain-NET | 0.91 | 0.95 | 0.67 | 38 | |
| | | | | | | | | | | | KPLS | 0.9 | 0.95 | 0.57 | 115 | |

| | | | | | | | | | | | | | | | | | |
|---|---|---|---|---|---|---|---|---|---|---|---|---|---|---|---|---|---|
| | | | | | | | | | | | SVR | 0.91 | 0.97 | 0.56 | 59 | | |
| | | | | | | | | | | | RF | 0.9 | 0.98 | 0.33 | 109 | | |
| **Affordable, web-based surgical skill training and evaluation tool[14]** | Gazi Islam, Kanav Kahol, Baoxin Li, Marshall Smith, Vimla L. Patel | 2016 | USA | Journal of Biomedical Informatics | 52 subjects (medical students and surgical residents) | LS | PT, SU, PC | VR | CV, NN | Custom scores | not available | | | | | | |
| **Tool Detection and Operative Skill Assessment in Surgical Videos Using Region-Based Convolutional Neural Networks[15]** | Amy Jin, Serena Yeung, Jeffrey Jopling, Jonathan Krause, Dan Azagury, Amy Jin, Serena Yeung, Jeffrey Jopling, Jonathan Krause, Dan Azagury, Arnold Milstein, and Li Fei-Fei | 2018 | USA | 2018 IEEE Winter Conference on Applications of Computer Vision (WACV) | unknown | Lap | Laparoscopic cholecystectomy | VR | CNN | unknown | qualitative results | | | | | | |
| **Machine Learning based Classification of Local Robotic Surgical Skills in a Training Tasks Set[16]** | L. Juarez-Villalobos, N. Hevia-Montiel, J. Perez-Gonzalez | 2021 | Mexico | 2021 43rd Annual International Conference of the IEEE Engineering in Medicine & Biology Society (EMBC) | 8 participants | RS | SU, NP, KT | KD (dV) | ML | binary (N, E) | Task | 10-fold cross validation | | | Final Test | | |
| | | | | | | | | | | | **Knot tying** | Accuracy | AUC-ROC | F1 Score | Accuracy | AUC-ROC | F1 Score |
| | | | | | | | | | | | KNN | 1.0 +- 0.0 | 1.0 +- 0.0 | 1.0 +- 0.0 | 1 | 1 | 1 |
| | | | | | | | | | | | RF (random forest) | 1.0 +- 0.0 | 1.0 +- 0.0 | 1.0 +- 0.0 | 1 | 1 | 1 |
| | | | | | | | | | | | SVM | 1.0 +- 0.0 | 1.0 +- 0.0 | 1.0 +- 0.0 | 1 | 1 | 1 |
| | | | | | | | | | | | **Needle-Passing** | | | | | | |
| | | | | | | | | | | | KNN | 1.0 +- 0.0 | 1.0 +- 0.0 | 1.0 +- 0.0 | 0.83 | 0.83 | 0.83 |
| | | | | | | | | | | | RF (random forest) | 0.95 +- 0.16 | 0.93+- 0.19 | 0.86+- 0.38 | 1 | 1 | 1 |
| | | | | | | | | | | | SVM | 1.0 +- 0.0 | 1.0 +- 0.0 | 1.0 +- 0.0 | 1 | 1 | 1 |
| | | | | | | | | | | | **Suturing** | | | | | | |
| | | | | | | | | | | | KNN | 1.0 +- 0.0 | 1.0 +- 0.0 | 1.0 +- 0.0 | 1 | 1 | 1 |
| | | | | | | | | | | | RF (random forest) | 0.97+- 0.11 | 0.97+- 0.09 | 0.96+- 0.12 | 1 | 1 | 1 |
| | | | | | | | | | | | SVM | 1.0 +- 0.0 | 1.0 +- 0.0 | 1.0 +- 0.0 | 1 | 1 | 1 |
| **High density optical neuroimaging predicts surgeons's subjective experience and skill levels[17]** | Hasan Onur Keles, Canberk Cengiz, Irem Demiral, Mehmet Mahir Ozmen, Ahmet Omurtag | 2021 | Turkey | PLoS ONE | 16 surgeons, 17 medical students | LS | PT, threading | NIRS (functional neuroimaging data) | ML | binary (student vs. Attending) | ~90% accuracy (precise results not available) | | | | | | |
| **Bidirectional long short-term memory for surgical skill classification of temporally segmented tasks[18]** | Jason D. Kelly, Ashley Petersen, Thomas S. Lendvay, Timothy M. Kowalewski | 2020 | USA | International Journal of Computer Assisted Radiology and Surgery | unknown | LS | 110 PT, 110 PC, 115 SU, 119 clipping | VR | CNN | binary (N, E) | | Accuracy | Novice-specific accuracy | Expert-specific accuracy | | | |
| | | | | | | | | | | | Suturing | 0.9688 | 1 | 0.9375 | | | |
| | | | | | | | | | | | Peg Transfer | 0.875 | 0.9375 | 0.8125 | | | |

| | | | | | | | | | | | | | | | | | | |
|---|---|---|---|---|---|---|---|---|---|---|---|---|---|---|---|---|---|---|
| | | | | | | | | | | | Cutting | 0.875 | 0.875 | 0.875 | | | | |
| | | | | | | | | | | | Clipping | 0.7333 | 0.6875 | 0.875 | | | | |
| **Evaluation of Deep Learning Models for Identifying Surgical Actions and Measuring Performance[19]** | Shuja Khalid, Mitchell Goldenberg, Teodor Grantcharov, Babak Taati, Frank Rudzicz | 2020 | Canada | JAMA Network Open | 8 | RS | SU, NP, KT | KD (dV) | CNN | N, I, E | Source | Data type | Scheme | Model | Metric | Novice | Intermediate | Expert |
| | | | | | | | | | | | Embedding analysis | Video | LOSO | bi-LSTM (attention) | Accuracy | 0.77 (0.14) | 0.77 (0.14) | 0.77 (0.14) |
| | | | | | | | | | | | | | | | Precision | 0.85 (0.09) | 0.67 (0.07) | 0.79 (0.12) |
| | | | | | | | | | | | | | | | Recall | 0.85 (0.05) | 0.69 (0.14) | 0.80 (0.13) |
| | | | | | | | | | | | | | | | F1 score | 0.85 (0.07) | 0.68 (0.10) | 0.79 (0.12) |
| | | | | | | | | | | | | | LOUO | Gated recurrent unit | Accuracy | 0.70 (0.21) | 0.70 (0.21) | 0.70 (0.21) |
| | | | | | | | | | | | | | | | Precision | 0.91 (0.05) | 0.48 (0.11) | 0.70 (0.15) |
| | | | | | | | | | | | | | | | Recall | 0.76 (0.08) | 0.67 (0.19) | 0.75 (0.11) |
| | | | | | | | | | | | | | | | F1 score | 0.83 (0.05) | 0.55 (0.12) | 0.72 (0.13) |
| | | | | | | | | | | | Key point representation analysis | Video | LOUO | Bi-LSTM | Accuracy | 0.73 (0.33) | 0.73 (0.33) | 0.73 (0.33) |
| | | | | | | | | | | | | | | | Precision | 1.00 (0) | 0.01 (0) | 1.00 (0) |
| | | | | | | | | | | | | | | | Recall | 0.47 (0.18) | 0.29 (0.11) | 1.00 (0) |
| | | | | | | | | | | | | | | | F1 score | 0.64 (0.25) | 0.02 (0.01) | 1.00 (0) |
| **Development and Validation of a 3-Dimensional Convolutional Neural Network for Automatic Surgical Skill Assessment Based on Spatiotemporal Video Analysis[20]** | Daichi Kitaguchi, Nobuyoshi Takeshita, Hiroki Matsuzaki, Takahiro Igaki, Hiro Hasegawa, Masaaki Ito | 2021 | Japan | JAMA Network Open | unknown | Lap | Laparoscopic colorectal surgery | VR | CNN | Endoscopic Surgical Skill Qualification System Score | Mean SD accuracy | | Overall | Medial mobilization | Lateral mobilization | IMA transection | Mesorectal transecion | |
| | | | | | | | | | | | | SD Accuracy | 75.0% (6.3%) | 73.00% | 74.30% | 83.80% | 68.90% | |
| **Sensor-based machine learning for workflow detection and as key to detect expert level in laparoscopic suturing and knot-tying[21]** | Karl-Friedrich Kowalewski, Carly R. Garrow, Mona W. Schmidt, Laura Benner, Beat P. Müller-Stich, Felix Nickel | 2019 | Germany | Surgical Endoscopy | 28 participants | LS | SU, KT | KD (s): Myoarmband | ML | Group classification (N, I, E) and OSATS score prediction | | ML algorithm type | Mean Error | r2 | | | | |
| | | | | | | | | | | | OSATS Score classification: | Decision forest | 4.45 ± 0.75 | − 0.2 ± 0.69 | | | | |
| | | | | | | | | | | | | Neural networks | 3.71 ± 0.64 | 0.03 ± 0.81 | | | | |
| | | | | | | | | | | | | Boosted decision tree | 4.43 ± 0.60 | 0.19 ± 0.46 | | | | |
| | | | | | | | | | | | Group classification | ML algorithm type | Accuracy | Precision | Recall | | | |
| | | | | | | | | | | | | Decision jungle | 0.62 | 0.43 | 0.43 | | | |
| | | | | | | | | | | | | Neural networks | 0.7 | 0.56 | 0.56 | | | |

| | | | | | | | | | | | | | | | |
|---|---|---|---|---|---|---|---|---|---|---|---|---|---|---|---|
| | | | | | | | | | | | | Support vector | 0.6 | 0.39 | 0.39 |
| | | | | | | | | | | | | Boosted decision tree | 0.66 | 0.56 | 0.56 |
| **Endoscopic Image-Based Skill Assessment in Robot-Assisted Minimally Invasive Surgery**[22] | Gábor Lajkó, Renáta Nagyné Elek, Tamás Haidegger | 2021 | Germany, Hungary, Austria | Sensors (Basel) | 8 participants | RS | SU, NP, KT | KD (dV) | CNN, CNN-LSTM (long short-term memory), ResNET (Residual Neural Network) | binary (N, E) | Efficacy | Suturing Task | Needle Passing | Knot tying | |
| | | | | | | | | | | | CNN | 0.8072 | 0.7966 | 0.8041 | |
| | | | | | | | | | | | CNN + LSTM | 0.8158 | 0.8319 | 0.8282 | |
| | | | | | | | | | | | ResNet | 0.8189 | 0.8423 | 0.8354 | |
| **Automation of surgical skill assessment using a three-stage machine learning algorithm**[23] | Joël L. Lavanchy, Joel Zindel, Kadir Kirtac, Isabell Twick, Enes Hosgor, Daniel Candinas & Guido Beldi | 2021 | Switzerland | Scientific Reports | 40 surgeons | Lap | Laparoscopic cholecystectomy | VR | CNN, ML | binary (good vs. Poor surgical skill) and skill level from 1-5 (with +/- 1 deviation) | Accuracy | Good vs. Poor surgical skill | skill level from 1-5 | | |
| | | | | | | | | | | | | 87 +/- 0.2% | 70 +/- 0.2% | | |
| **Artificial Neural Network for Laparoscopic Skills Classification Using Motion Signals from Apple Watch**[24] | Rubbermaid Laverde, Claudia Rueda, Lusvin Amado, David Rojas, and Miguel Altuve | 2018 | Colombia | 2018 40th Annual International Conference of the IEEE Engineering in Medicine and Biology Society (EMBC) | 7 volunteers without experience in laparoscopy | LS | PT | KD (s): Apple watch | ANN | N, I, E | Average iteration (of 5) | F1 low | F1 intermediate | F1 high | F1 score |
| | | | | | | | | | | | | 85.14% | 91.21% | 81.97% | 86.11% |
| **Surgeon Technical Skill Assessment using Computer Vision based Analysis**[25] | Hei Law, Khurshid Ghani, Jia Deng | 2017 | USA | Proceedings of the 2nd Machine Learning for Healthcare Conference | 12 surgeons | Rob | Robotic prostatectomy | VR | CV, ML (SVM), NN | binary (good vs. poor surgical skill) | | Accuracy | | | |
| | | | | | | | | | | | right hand only | 83.33% | | | |
| | | | | | | | | | | | right + left hand annotations | 91.67% | | | |
| **Evaluation of Surgical Skills during Robotic Surgery by Deep Learning-Based Multiple Surgical Instrument Tracking in Training and Actual Operations**[26] | Dongheon Lee, Hyeong Won Yu, Hyungju Kwon, Hyoun-Joong Kong, Kyu Eun Lee and Hee Chan Kim | 2020 | Korea | Journal of Clinical Medicine | unknown | RS, Rob | Robotic thyroid surgery (DaVinci) performed on patients with thyroid cancer, bilateral axillo-breast approach (BABA) training model | VR | DL, CNN | N, I, E | Accuracy | OSATS | GEARS | | |
| | | | | | | | | | | | Linear classifier | 58% | 67% | | |
| | | | | | | | | | | | SVM | 75% | 67% | | |
| | | | | | | | | | | | RF (random forest) | 83% | 83% | | |
| **Clearness of operating field: a surrogate for surgical skills on in vivo clinical data**[27] | Daochang Liu, Tingting Jiang, Yizhou Wang, Rulin Miao, Fei Shan, Ziyu Li | 2020 | China | International Journal of Computer Assisted Radiology and Surgery | unknown | Lap | Laparoscopic gastrectomy | VR | NN | modified OSATS score: 14 skill metrics (1-5pts. Each), OPS: 7 metrics (1-5pts each) | | Method | % SROCC (Std.) | % PLOCC (Std.) | |
| | | | | | | | | | | | OTS =overall technical skills | Direct (automated, L(both)) | 49.4 (0.5) | 53.5 (1.3) | |

| | | | | | | | | | | | | | | | | | | |
|---|---|---|---|---|---|---|---|---|---|---|---|---|---|---|---|---|---|---|
| | | | | | | | | | | | | Surrogate (automated, L(both)) | 59.5 (0.4) | 61.5 (0.2) | | | | |
| | | | | | | | | | | | OPS = overall procedural skills | Direct (automated, L(both)) | 25.5 (1.3) | 25.0 (1.9) | | | | |
| | | | | | | | | | | | | Surrogate (automated, L(both)) | 41.6 (0.4) | 42.6 (0.5) | | | | |
| **Towards Unified Surgical Skill Assessment[28]** | Daochang Liu, Qiyue Li, Tingting Jiang, Yizhou Wang, Rulin Miao, Fei Shan, Ziyu Li | 2021 | China | IEEE/CVF Conference on Computer Vision and Pattern Recognition (CVPR) | 8 participants/unknown | RS, Lap | SU, NP, KT / 20 laparoscopic videos of in vivo surgeries for gastric cancer | VR | ML | modified OSATS score | SROCC | | Suturing | Needle Passing | Knot Tying | Avg. | | |
| | | | | | | | | | | | Clinical Dataset | 0.565 | | | | | | |
| | | | | | | | | | | | JIGSAWS DATASET | | 0.83 | 0.76 | 0.82 | 0.8 | | |
| **An objective approach to evaluate novice robotic surgeons using a combination of kinematics and stepwise cumulative sum (CUSUM) analyses[29]** | William B. Lyman, Michael J. Passeri, Keith Murphy, Imran A. Siddiqui, Adeel S. Khan, David A. Iannitti, John B. Martinie, Erin H. Baker, Dionisios Vrochides | 2021 | USA | Surgical Endoscopy | 2 hepatopancreatobiliary surgery (HPB) fellows | RS | 40 robotic assisted hepaticojejunostomy reconstruction | KD (dV) | ML | binary (N, I) | Accuracy | 89.30% | | | | | | |
| **Surgical skill levels: Classification and analysis using deep neural network model and motion signals[30]** | Xuan Anh Nguyen, Damir Ljuhar, Maurizio Pacilli, Ramesh Mark Nataraja, Sunita Chauhan | 2019 | Australia | Computer Methods and Programs in Biomedicine | 8 participants | RS | SU, NP, KT | KD (dV) | DNN | N, I, E | JIGSAWS DATASET (average classification results %) | Suturing | Needle passing | knot tying | | | | |
| | | | | | | | | | | | CNN-LSTM | 97.2 | 97.3 | 91.5 | | | | |
| | | | | | | | | | | | CNN-LSTM + SENET | 98.3 | 97.8 | 94.7 | | | | |
| | | | | | | | | | | | CNN-LSTM + SENET + Restart | 98.4 | 98.4 | 94.8 | | | | |
| **Automatically rating trainee skill at a pediatric laparoscopic suturing task[31]** | Yousi A. Oquendo, Elijah W. Riddle, Dennis Hiller, Thane A. Blinman, Katherine J. Kuchenbecker | 2018 | USA | Surgical Endoscopy | 32 participants (med students - fellows) | LS | SU | KD (s): magnetic sensors | ML | OSATS score (summed scores and rounded average scores) | Scoring Performance | Summed Scores | | | | | | |
| | | | | | | | | | | | | Plus/minus 2 Accuracy | Plus/minus 4 Accuracy | Correlation | | | | |
| | | | | | | | | | | | TMVG (model that used all sensor data streams) | 0.71 | 0.89 | 0.85 | | | | |
| **Objective classification of psychomotor laparoscopic skills of surgeons based on three different approaches[32]** | Fernaando Pérez-Escamirosa, Antonio Alarcón-Paredes, Gustavo Adolfo | 2019 | Mexico | International Journal of Computer Assisted Radiology and Surgery | 43 participants (med students - experienced surgeons) | LS | PC, PT, SU | VR | ML (neural network based (RBFNet), lazy learner | binary (experienced, non-experienced) | Task | Hold out (75–25) | | | | | | |
| | | | | | | | | | | | | Classifier | Accuracy (%) | RMSE | Sensitivity (%) | Specificity (%) | AUC | F1-Score |
| | | | | | | | | | | | Peg transfer | RBFNets | 81.82 | 0.387 | 81.8 | 72.3 | 0.92 | 0.75 |

| | | | | | | | | | | | | | | | | | | |
|---|---|---|---|---|---|---|---|---|---|---|---|---|---|---|---|---|---|---|
| | Alonso-Silverio, Ignacio Oropesa, Oscar Camacho-Nieto, Daniel Lorias-Espinoza, Arturo Minor-Martínez | | | | | | | | based on distance computation (K-star), tree-based classifier (random forest) | | | K* | 90.91 | 0.305 | 90.9 | 75.8 | 0.92 | 0.8 |
| | | | | | | | | | | | | RF | 81.82 | 0.337 | 81.8 | 72.3 | 0.92 | 0.75 |
| | | | | | | | | | | | | Avg | 84.85 | 0.343 | 84.83 | 73.46 | 0.92 | 0.77 |
| | | | | | | | | | | | Pattern cutting | RBFNets | 90.01 | 0.304 | 90.9 | 75.8 | 0.96 | 0.8 |
| | | | | | | | | | | | | K* | 98.18 | 0.066 | 100 | 97.55 | 0.99 | 0.92 |
| | | | | | | | | | | | | RF | 81.82 | 0.331 | 81.8 | 72.3 | 0.92 | 0.75 |
| | | | | | | | | | | | | Avg | 90.61 | 0.317 | 90.9 | 82.7 | 0.96 | 0.82 |
| | | | | | | | | | | | Intracorporeal knot suture | RBFNets | 90.91 | 0.302 | 90.9 | 75.8 | 0.96 | 0.8 |
| | | | | | | | | | | | | K* | 90.91 | 0.302 | 90.9 | 96.6 | 0.96 | 0.86 |
| | | | | | | | | | | | | RF | 90.91 | 0.245 | 90.9 | 75.8 | 0.96 | 0.8 |
| | | | | | | | | | | | | Avg | 90.91 | 0.283 | 90.9 | 82.73 | 0.96 | 0.82 |
| | | | | | | | | | | | Task | Leave-one-out cross-validation | | | | | | |
| | | | | | | | | | | | | Classifier | Accuracy (%) | RMSE | Sensitivity (%) | Specificity (%) | AUC | F1-Score |
| | | | | | | | | | | | Peg transfer | RBFNets | 90.7 | 0.239 | 90.7 | 83.2 | 0.94 | 0.8 |
| | | | | | | | | | | | | K* | 86.05 | 0.336 | 86 | 74.9 | 0.96 | 0.86 |
| | | | | | | | | | | | | RF | 81.4 | 0.324 | 81.4 | 66.5 | 0.9 | 0.76 |
| | | | | | | | | | | | | Avg | 86.05 | 0.299 | 86.03 | 74.86 | 0.93 | 0.81 |
| | | | | | | | | | | | Pattern cutting | RBFNets | 86.05 | 0.329 | 86 | 81.8 | 0.95 | 0.73 |
| | | | | | | | | | | | | K* | 90.7 | 0.274 | 90.7 | 97.2 | 0.99 | 0.83 |
| | | | | | | | | | | | | RF | 76.74 | 0.341 | 76.7 | 58.1 | 0.89 | 0.67 |
| | | | | | | | | | | | | Avg | 84.5 | 0.314 | 84.46 | 79.03 | 0.94 | 0.74 |
| | | | | | | | | | | | Intracorporeal knot suture | RBFNets | 93.02 | 0.273 | 93 | 77 | 0.9 | 0.86 |
| | | | | | | | | | | | | K* | 97.67 | 0.153 | 97.7 | 93 | 0.99 | 0.95 |
| | | | | | | | | | | | | RF | 93.02 | 0.197 | 93 | 90.9 | 0.99 | 0.86 |
| | | | | | | | | | | | | Avg | 94.57 | 0.207 | 94.56 | 86.96 | 0.96 | 0.89 |
| **Surgical Skill Evaluation From Robot-Assisted Surgery Recordings**[33] | Abed Soleymani, Ali Akbar Sadat Asl, Mojtaba Yeganejou, Scott Dick, Mahdi Tavakoli, Xingyu Li | 2021 | Canada | 2021 International Symposium on Medical Robotics (ISMR) | 8 participants | RS | SU, NP, KT | VR | DL | N, I, E | Accuracy | 97.27 +/- 2.35 | | | | | | |

| | | | | | | | | | | | | | | | | | | | |
|---|---|---|---|---|---|---|---|---|---|---|---|---|---|---|---|---|---|---|---|
| **Feasibility of an AI-Based Measure of the Hand Motions of Expert and Novice Surgeons[34]** | Munenori Uemura , Morimasa Tomikawa, Tiejun Miao, Ryota Souzaki, Satoshi Ieiri, Tomohiko Akahoshi, Alan K. Lefor, Makoto Hashizume | 2018 | Japan | Computational and Mathematical Methods in Medicine | 67 surgeons | LS | SU | KD (s): magnetic sensors | NN | Novice, Expert | correctly distinguished | 79% of the participants | | | | | | | |
| **Evaluating robotic-assisted surgery training videos with multi-task convolutional neural networks[35]** | Yihao Wang, Jessica Dai, Tara N. Morgan, Mohamed Elsaied, Alaina Garbens, Xingming Qu, Ryan Steinberg, Jeffrey Gahan, Eric C. Larson | 2021 | USA | Journal of Robotic Surgery | 18 surgeons | RS | SU (artificial urinary tissue in the final step of a prostatectomy) | VR | CNN | GEARS scores (1-5)// expert >25, intermediate 20–25, and novice <20. | GEARS Score +/- 1 point | 86.10% | | | | | | | |
| | | | | | | | | | | | GEARS Score +/- 2 points | 1 | | | | | | | |
| | | | | | | | | | | | | Depth Perception | Bi-manual Dexterity | Efficiency | Force Sens. | Autonomy | Robotic control | | |
| | | | | | | | | | | | Matching | 94.40% | 77.80% | 94.40% | 83.30% | 72.20% | 88.90% | | |
| | | | | | | | | | | | **Expert vs. Intermediate vs. Novice** | | | | | | | | |
| | | | | | | | | | | | Classified correctly | 83.50% | | | | | | | |
| | | | | | | | | | | | Novice | 100% (11/11) | | | | | | | |
| | | | | | | | | | | | Intermediate | 80% (4/5) | | | | | | | |
| | | | | | | | | | | | expert | 0% (0/2) | | | | | | | |
| **Deep learning with convolutional neural network for objective skill evaluation in robot-assisted surgery[36]** | Ziheng Wang, Ann Majewicz Fey | 2018 | USA | International Journal of Computer Assisted Radiology and Surgery | 8 participants | RS | SU, NP, KT | KD (dV) | DL, CNN | N, I, E (self-proclaimed > practice hours vs. GRS-based) | Accuracy | self-proclaimed | GRS based | | | | | | |
| | | | | | | | | | | | Suturing | 93.4 | 92.5 | | | | | | |
| | | | | | | | | | | | Needle passing | 89.8 | 95.4 | | | | | | |
| | | | | | | | | | | | Knot tying | 84.9 | 91.3 | | | | | | |
| **SATR-DL: Improving Surgical Skill Assessment and Task Recognition in Robot-assisted Surgery with Deep Neural Networks[37]** | Ziheng Wang, Ann Majewicz Fey | 2018 | USA | 2018 40th Annual International Conference of the IEEE Engineering in Medicine and Biology Society (EMBC) | 8 participants | RS | SU, NP, KT | KD (dV) | DNN | N, I, E | | Interval-level classification | | | | trial-level classification | | | |
| | | | | | | | | | | | | Precision | Recall | F1-score | Overall Accuracy | Precision | Recall | F1-score | Overall Accuracy |
| | | | | | | | | | | | Novice | 0.94 | 0.96 | 0.95 | 0.92 | 0.95 | 0.98 | 0.97 | 0.966 |
| | | | | | | | | | | | Intermediate | 0.88 | 0.77 | 0.82 | | 1 | 0.9 | 0.95 | |
| | | | | | | | | | | | Expert | 0.9 | 0.95 | 0.93 | | 0.97 | 1 | 0.98 | |

Abbreviations: n/a = not available, LS = laparoscopic simulator, Lap = laparoscopic surgery, RS = robotic simulator, Rob = robotic surgery, PC = pattern cutting, SU = suturing, NP = needle-passing, KT = knot-tying, PT = peg transfer, VR = video recordings, KD (dv) = kinematic data collected by daVinci systems, fNIRS = functional near-infrared spectroscopy, KD (s) = kinematic data collected by external sensors, ANN = artificial neural network, CV = computer vision, CNN = convolutional neural network, DL = deep learning, ML = machine learning, N = novice, I = intermediate, E = expert, PLACE = Pelvic Lymphadenectomy Assessment and Completion Evaluation, GEARS = Global Evaluative Assessments of Robotic Skills, FLS = Fundamentals of Laparoscopic Surgery, ESSQS = Endoscopic Surgical Skill Qualification System, OSATS = Objective Structured Assessment of Technical Skills

Table S3: Risk of bias assessment of included studies

| Study | Population | Index test | Reference standard | Flow and timing | Data management |
|---|---|---|---|---|---|
| | Was the study population described? | Was cross-validation or external validation performed? | Who performed the annotation? Performance validation? | Did all participants receive the same reference standard? Were all patients included in the analysis | Was ethical approval specified? |
| Alonso-Silverio et al.[1] | ✓ | ✓ | ✗ | ✓ | ✗ |
| Anh et al.[2] | ✓ | ✗ | ✗ | ✓ | ✗ |
| Baghdadi et al.[3] | ✗ | ✗ | ✗ | ✓ | ✗ |
| Benmansour et al.[4] | ✗ | ✗ | ✗ | ✗ | ✗ |
| Brown et al.[5] | ✓ | ✓ | ✓ | ✓ | ✓ |
| Castro et al.[6] | ✓ | ✓ | ✗ | ✓ | ✗ |
| Fard et al.[7] | ✗ | ✓ | ✗ | ✓ | ✗ |
| Fathabadi et al.[8] | ✗ | ✓ | ✗ | ✗ | ✗ |
| Fawaz et al.[9] | ✓ | ✓ | ✗ | ✓ | ✗ |
| Forestier et al.[10] | ✓ | ✓ | ✗ | ✓ | ✗ |
| French et al.[11] | ✓ | ✓ | ✗ | ✗ | ✓ |
| Funke et al.[12] | ✓ | ✓ | ✗ | ✓ | ✓ |
| Gao et al.[13] | ✓ | ✓ | ✗ | ✓ | ✓ |
| Islam et al.[14] | ✗ | ✗ | ✓ | ✓ | ✗ |
| Jin et al.[15] | ✗ | ✗ | ✓ | ✓ | ✗ |
| Juarez-Villalobos et al.[16] | ✓ | ✓ | ✗ | ✓ | ✗ |
| Keles et al.[17] | ✓ | ✓ | ✗ | ✓ | ✓ |
| Kelly et al.[18] | ✓ | ✓ | ✓ | ✓ | ✓ |
| Khalid et al.[19] | ✓ | ✓ | ✗ | ✓ | ✓ |
| Kitaguchi et al.[20] | ✓ | ✓ | ✓ | ✓ | ✓ |
| Kowalewski et al.[21] | ✓ | ✓ | ✓ | ✓ | ✓ |

| | | | | | |
|---|---|---|---|---|---|
| Lajkó et al.[22] | ✘ | ✓ | ✘ | ✓ | ✘ |
| Lavanchy et al.[23] | ✓ | ✓ | ✓ | ✓ | ✓ |
| Laverde et al.[24] | ✓ | ✓ | ✘ | ✓ | ✓ |
| Law et al.[25] | ✓ | ✓ | ✓ | ✓ | ✘ |
| Lee et al.[26] | ✘ | ✓ | ✘ | ✓ | ✘ |
| Liu et al.[27] | ✓ | ✘ | ✓ | ✓ | ✓ |
| Liu et al.[28] | ✓ | ✓ | ✓ | ✓ | ✘ |
| Lyman et al.[29] | ✓ | ✘ | ✘ | ✘ | ✘ |
| Nguyen et al.[30] | ✓ | ✓ | ✘ | ✓ | ✘ |
| Oquendo et al.[31] | ✓ | ✓ | ✓ | ✓ | ✓ |
| Pérez-Escamirosa et al.[32] | ✓ | ✓ | ✘ | ✓ | ✓ |
| Soleymani et al.[33] | ✓ | ✓ | ✘ | ✓ | ✘ |
| Uemura et al.[34] | ✓ | ✓ | ✘ | ✘ | ✘ |
| Wang Y. et al.[35] | ✘ | ✓ | ✘ | ✓ | ✓ |
| Wang Z. et al.[36] | ✓ | ✓ | ✘ | ✓ | ✓ |
| Wang Z. et al.[37] | ✘ | ✓ | ✘ | ✓ | ✘ |

✓ = low risk of bias, ✘ = high risk of bias
